# Age-related nonlinear trajectories of abdominal organ volumes on CT: a longitudinal study

**DOI:** 10.64898/2026.05.06.26352299

**Authors:** Yukihiro Nomura, Shouhei Hanaoka, Takahiro Nakao, Yosuke Yamagishi, Tomohiro Kikuchi, Yuki Sonoda, Soichiro Miki, Koji Oba, Takeharu Yoshikawa, Osamu Abe

## Abstract

**Objectives:** To characterize longitudinal age-related changes in abdominal organ volumes using CT volumetry and to model nonlinear trajectories across multiple organs.

**Materials & Methods:** This retrospective single-center study included adults who underwent whole-body screening low-dose CT between 2006 and 2017. Subjects with at least eight examinations during a follow-up period of at least 78 months were included. After applying exclusion criteria, 700 participants with 6,739 CT series were analyzed. Non-contrast CT images were processed using automated organ segmentation, and volumes of the liver, pancreas, spleen, and kidneys were quantified. Longitudinal changes were modeled using generalized additive mixed models with sex-specific smooth functions of age and subject-level random effects. Age-dependent rates of change were estimated from model derivatives.

**Results:** A total of 700 participants (mean age, 56.9 ± 9.8 years, 29.6% women) were evaluated. Liver, pancreas, and kidney volumes showed mild increases or plateaued at approximately 40–60 years of age, depending on the organ, and were followed by gradual declines with advancing age, whereas splenic volume showed a progressive decrease across the age range. These patterns showed nonlinear age dependence. The transition from positive to negative change rates tended to occur earlier in women than in men for several organs, particularly the liver and kidneys.

**Conclusion:** Longitudinal CT analysis demonstrated nonlinear age-related changes in abdominal organ volumes, with organ-specific trajectories and sex-related differences in the timing and magnitude of volume changes.

**Question:** How do abdominal organ volumes change longitudinally with age, and can their trajectories be characterized for each organ?

**Findings:** Longitudinal CT analysis demonstrated nonlinear, organ-specific volume trajectories, with transitions from stability to decline around 40–60 years and earlier transitions in women than men.

**Clinical Relevance:** Longitudinal reference patterns of abdominal organ volumes on CT improve the interpretation of age-related changes and support more accurate differentiation between physiological variation and disease-related volume alterations.

## Introduction

Organ volume measurements from CT are increasingly recognized as imaging biomarkers associated with functional status, disease progression, and clinical outcomes [1–3], including associations with renal function decline, portal hemodynamics, and metabolic dysfunction. Aging is recognized as a nonlinear process across biological systems, with studies suggesting age-dependent, nonlinear changes in molecular profiles [4, 5]. At the imaging level, age-related changes in organ structure and body composition are not well captured by simple linear models, as shown in volumetric imaging studies of brain structure [6] and body composition across the lifespan [7].

Recent advances in deep learning-based whole-body organ segmentation, such as TotalSegmentator [8] and VIBESegmentator [9], have enabled automated and large-scale quantification of organ volumes. This progress has facilitated population-level and multi-organ analyses in large imaging cohorts, as demonstrated in recent CT studies [10, 11]. Among these, Kikuchi et al. [10] established age- and sex-specific distributions of abdominal organ volumes, whereas Wachinger et al. [11] demonstrated organ-specific patterns of age-related changes across multiple organs using cross-sectional and longitudinal analyses. However, the datasets used in these studies were primarily derived from CT acquired for clinical indications, and many pathological organs appear to have been included in the analyses. Kikuchi’s study employed a cross-sectional design, and even in Wachinger’s study, longitudinal assessment was limited because of substantial variability in the number of follow-up examinations and the duration of follow-up. A limitation of cross-sectional designs is that it cannot distinguish true within-person age-related changes from generational differences caused by environmental factors. These factors may have prevented the previous studies from capturing subtle age-related changes in healthy adults.

In this study, we aimed to characterize longitudinal changes in abdominal organ volumes using a large cohort of low-dose CT scans from an annual whole-body medical screening program, and to model age-related trajectories across multiple organs using a flexible statistical framework that captures nonlinear effects and intra-individual variability.

## Materials and methods

### Study population

This retrospective study was approved by the Research Ethics Committee of the Faculty of Medicine of the University of Tokyo (serial number: 1494-(22), approval date: 22 September 2006, last renewal date: 18 September 2025). The participants in this study comprised adults who visited The University of Tokyo Hospital for an annual whole-body medical screening program between November 2006 and November 2017. In this program, all participants aged 40 years or older underwent low-dose whole-body CT (from the neck to the pelvis) unless there were contraindications or they declined the examination. All subjects provided written informed consent for their medical images to be used for research purposes. The CT images were acquired using two GE LightSpeed CT scanners (GE Healthcare, Waukesha, WI) with the following parameters: number of detector rows, 16; tube voltage, 120 kVp; tube current, 30–210 mA (automatic exposure control); noise index, 25; rotation time, 0.5 s; moving table speed, 70 mm/s; body filter, soft; reconstruction slice thickness and interval, 1.25 mm; field of view, 500 mm; matrix size, 512 × 512 pixels; pixel spacing, 0.98 mm. All images underwent independent double reading by two board-certified radiologists, followed by consensus finalization of per-organ findings and institutionally defined 1-to-5 significance codes in a structured reporting system.

A total of 7,837 subjects with 23,712 CT series were initially collected. The longitudinal analysis dataset was constructed using predefined inclusion and exclusion criteria. Inclusion criteria were: (1) at least eight screening visits during the study period and (2) an interval of at least 78 months between the first and last examinations, yielding 746 subjects with 7,175 CT series. These criteria corresponded to approximately 11–12-month follow-up intervals, resulting in at least eight longitudinal time points per participant (see Supplementary Material, Fig. S2), and ensured sufficient follow-up duration and sample size for stable estimation of age-related trajectories. Exclusion criteria were: (1) cases with clinically significant abnormalities in the target organs (liver, pancreas, spleen, or kidneys; determined by a significant code ≥ 4) requiring additional evaluation or close follow-up, (2) prior surgical resection, including partial or total removal, of any of the target organs, and (3) congenital or anatomical abnormalities, including renal agenesis and other major structural variations such as situs inversus. After exclusion, the final longitudinal dataset comprised 700 subjects with 6,739 CT series (Fig. 1).

**Fig. 1.**
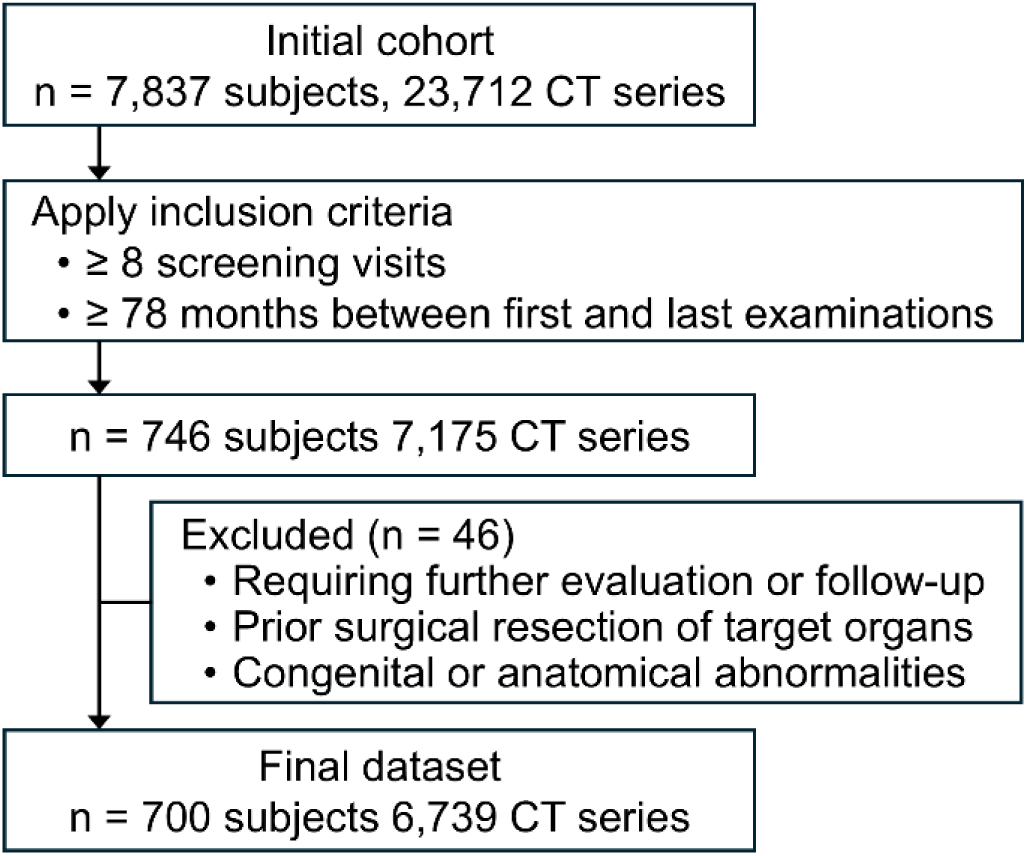
Flowchart of the subject selection process for the longitudinal analysis cohort.

### Image Processing

All CT series included in the study were processed using TotalSegmentator (version 2.10.0) for automated organ segmentation [8]. The processing pipeline was executed on an NVIDIA DGX A100 server equipped with two AMD Rome 7742 processors (AMD Inc., Santa Clara, CA), 2 TB of memory, and eight graphics processing units (GPUs) (A100 with 40 GB of memory, NVIDIA, Santa Clara, CA). All series were processed in parallel, with one series assigned to each GPU. The liver, pancreas, spleen, right kidney, and left kidney were analyzed as target organs. The segmented volume was calculated for each organ using custom Python scripts. The segmentation performance of TotalSegmentator was validated in a subset of the study cohort using manually annotated reference standards (Supplementary Material, Section S1).

### Statistical analysis

Prior to statistical analysis, outliers in organ volume measurements, which may reflect segmentation errors, were identified and removed using robust a statistical filtering approach adapted from a previous study [11]. Briefly, outliers were detected based on the distribution of organ volumes using robust statistical measures. Details of the procedure are provided in the Supplementary Material (Section S2).

For longitudinal analysis, age-related changes in organ volume were assessed using a generalized additive mixed model (GAMM) [12, 13]. Log-transformed organ volume was used as the outcome variable, with sex included as a fixed effect. To allow for potentially nonlinear age effects, age was modeled using sex-specific smooth functions estimated with penalized thin plate regression splines. Repeated measurements within the same individual were accounted for by including subject-specific random intercepts and random slopes for age. The model was specified as:

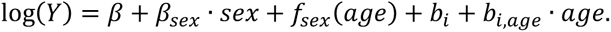

Here, 𝑓*_sex_*(*age*) represents a smooth age function for each sex, while 𝑏*_𝑖_* and 𝑏*_𝑖,age_* represent patient-specific random intercepts and random slopes, respectively. Smooth terms were fitted with penalized thin plate regression splines (*k* = 5, selected to avoid overfitting) with shrinkage selection to reduce overfitting. Age-dependent local change rates were then obtained from the first derivative of the fitted smooth age function and expressed as percentage change per year, with 95% confidence intervals derived from the corresponding standard errors.

As a secondary analysis, the previously reported baseline age–time model [11] was fitted. Certain covariates, including scanner-related and contrast-related terms, were “omitted because a single scanner model and non-contrast CT were used (see Supplementary Material, Section S3 for details).

All statistical analyses were performed using R version 4.5.3 (The R Project for Statistical Computing, Vienna, Austria), and GAMMs were fitted using the mgcv package version 1.9-4. Statistical interpretation was based on model-derived trajectories and their variability rather than formal hypothesis testing.

## Results

### Study cohort

The final longitudinal dataset comprised 700 participants (Fig. 1). The mean age at the first scan was 56.9 ± 9.8 years, and 29.6% of the participants were women. The number of screening visits ranged from 8 to 13 (Table 1). The distribution of examination intervals is shown in the Supplementary Material (Fig. S2). The majority of participants underwent follow-up examinations at approximately annual (11–12-month) intervals.

**Table 1.**
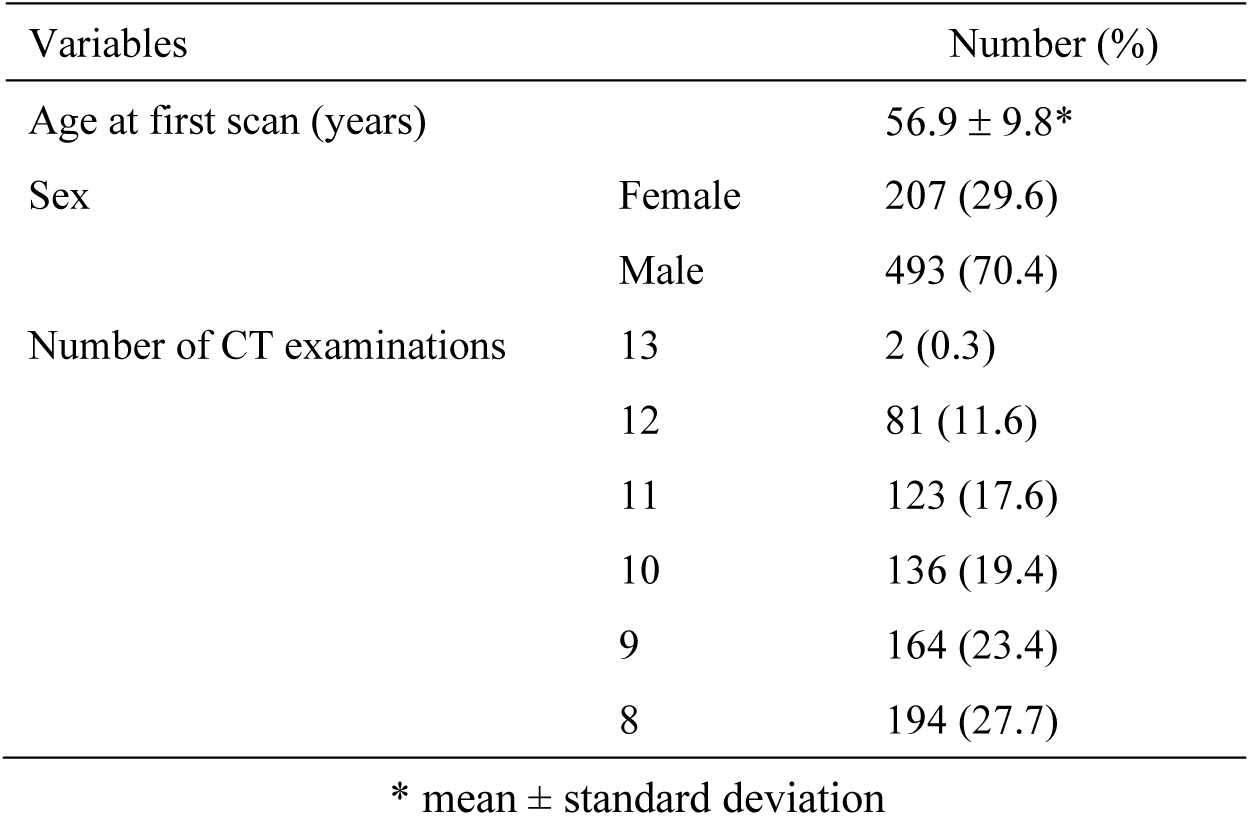
Demographic characteristics of the study cohort (n = 700).

| Variables |  | Number (%) |
| --- | --- | --- |
| Age at first scan (years) |  | 56.9 ± 9.8* |
| Sex | Female | 207 (29.6) |
|  | Male | 493 (70.4) |
| Number of CT examinations | 13 | 2 (0.3) |
|  | 12 | 81 (11.6) |
|  | 11 | 123 (17.6) |
|  | 10 | 136 (19.4) |
|  | 9 | 164 (23.4) |
|  | 8 | 194 (27.7) |

\* mean ± standard deviation

### Segmentation performance

The segmentation performance of TotalSegmentator was evaluated in a subset of the study cohort (Supplementary Material, Section S1). High agreement with manual annotations was observed for most organs, with Dice coefficients exceeding 0.90 for the liver, spleen, and kidneys, whereas the accuracy was lower for the pancreas.

### Longitudinal analysis

Figure 2 shows age-related changes in organ volume estimated using the GAMM. Liver, pancreas, and kidney volumes showed mild increases or plateaus at approximately 40–60 years of age, followed by gradual declines with increasing age, whereas splenic volume showed a progressive decrease across the age range. This overall age-related pattern was observed in both sexes, with consistently larger estimated organ volumes in males than in females. The results of the baseline age–time model applied to the same dataset are shown in Supplementary Fig. S1.

**Fig. 2.**
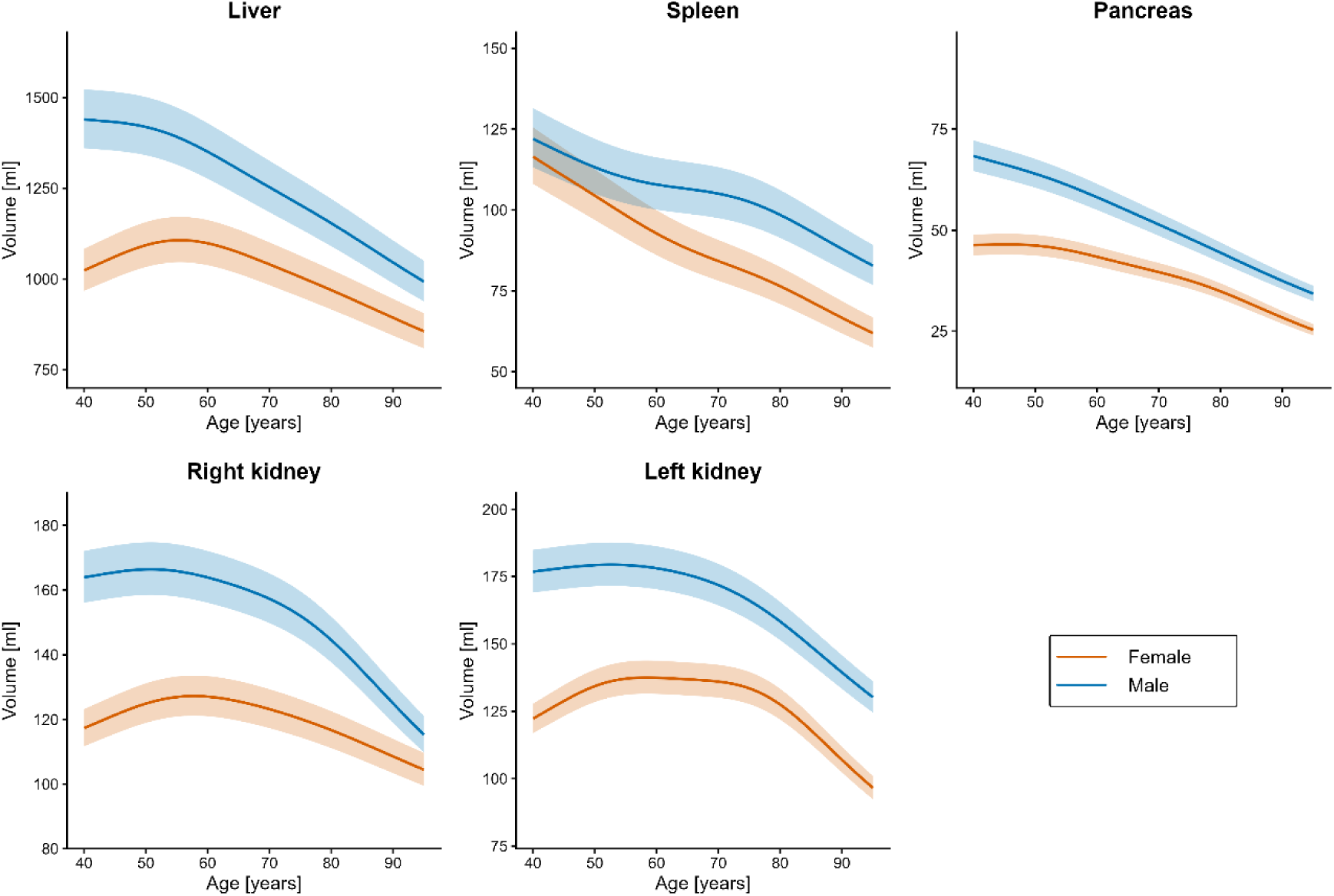
Visualization of generalized additive mixed model fits for organ volumes from the longitudinal analysis. Curves show estimated organ volumes as a function of age for female participants (red) and male participants (blue). Shaded ribbons indicate model-based 25th–75th percentile bands.

Figure 3 shows age-related annual percentage rates of change in organ volume derived from the trajectories shown in Fig. 2. In both sexes, liver, pancreas, and kidney volumes exhibited positive or near-zero change rates at earlier ages (approximately 40–60 years of age, depending on the organ), followed by progressively negative change rates with increasing age. In contrast, splenic volume showed generally negative change rates across most of the age range. The transition from positive to negative change rates tended to occur earlier in females than in males for several organs, particularly the liver and kidneys.

**Fig. 3.**
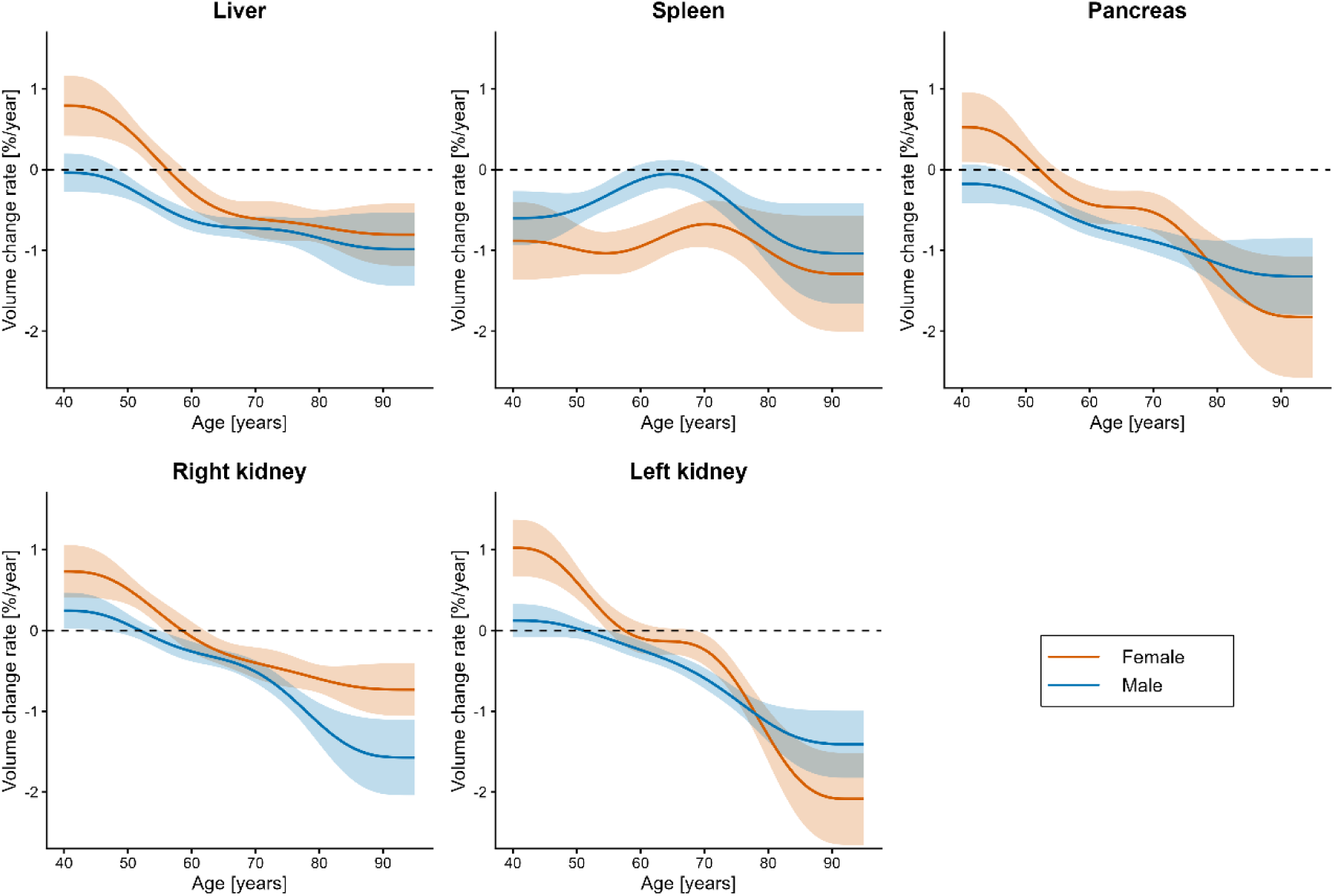
Age-related changes in organ volume change rates estimated using the generalized additive mixed model. Curves show the estimated annual percentage change in organ volume (%/year) as a function of age for female participants (red) and male participants (blue). Shaded ribbons indicate 95% confidence intervals, and dashed horizontal lines represent zero change.

## Discussion

This study provides a longitudinal characterization of age-related abdominal organ volume trajectories using large-scale serial CT data. To our knowledge, few studies have provided fully longitudinal assessments of abdominal organ volume changes across multiple organs under relatively physiological conditions. By analyzing 700 adults with at least 78 months of follow-up, as well as excluding target organs with clinically significant abnormalities, we were able to capture age-related volume changes under conditions closer to physiological aging. Liver, pancreas, and kidney volumes showed a mild increase or plateau at approximately 40–60 years of age, whereas splenic volume exhibited a more consistent decrease across the age range. While overall patterns were similar between sexes, differences were suggested in the timing and magnitude of these volume changes.

The overall patterns observed in this study were in agreement with those reported in previous studies. In particular, the age- and sex-specific distributions of abdominal organ volumes were comparable to those reported by Kikuchi et al. [10], who analyzed a large Japanese cohort, supporting the consistency of these findings within similar populations. The observed trajectories were characterized by relatively stable or mildly increasing volumes up to approximately 60 years of age, followed by gradual declines thereafter. Sex-related differences were consistently observed, with larger organ volumes in men than in women and differences in the timing of these changes, consistent with previous CT-based studies [14]. The age-related trajectories observed in this study are compatible with known physiological aging processes [15–17]. In addition, the overall trajectory shapes were similar to those obtained using a baseline age–time model, originally described by Wachinger et al. [11], when applied to the same dataset (Fig. S1). This consistency suggests that the observed age-related changes in organ volumes are robust features that can be captured across different modeling approaches, while the present model enables a more direct characterization of continuous age-related trajectories. In addition, dense longitudinal data with repeated examinations enabled more reliable characterization of intra-individual volume changes.

The age-related rates of organ volume change observed in this study provide additional insight into the dynamics of organ volume changes. While the present study quantified relative changes as percentage change per year based on longitudinal data, Wachinger et al. reported absolute changes in mL/year derived from cross-sectional age-based estimates rather than within-individual longitudinal changes using generalized additive models for location, scale, and shape [11]. Although these measures are not directly comparable, both approaches indicate gradual age-related declines in abdominal organ volumes. Similarly, Kimura et al. reported an approximately 0.95% annual decrease in liver volume using a linear model [18], supporting the magnitude of relative decline observed in the present study.

The age-related changes in organ volume and their rates of change identified in this study may serve as structural imaging biomarkers reflecting functional status, disease progression, and clinical outcomes [1–3]. In addition, these longitudinal reference trajectories may provide a basis for biological age estimation, in which deviations from expected age-related patterns are interpreted as indicators of accelerated or delayed aging, as demonstrated in recent imaging-based biological age studies [19].

This study has several limitations. First, the generalizability of our findings may be limited because this study was conducted at a single institution, and the participants were predominantly from a single ethnic background. In addition, while the use of standardized screening CT is a strength of this study, the cohort may have been relatively health-conscious, and the possibility of selection bias cannot be fully ruled out. Second, the study population was restricted to individuals aged 40 years and older, and the applicability of the findings to younger populations remains uncertain. Third, organ volumes were not adjusted for body size. While such normalization may facilitate inter-individual comparisons, it may also obscure clinically relevant changes when body size and organ volume are interrelated. Fourth, this study focused on abdominal organs, whereas previous studies have analyzed a broader range of organs. Fifth, the segmentation performance for the pancreas was lower than that for the other organs. In a subset of the study cohort, the Dice similarity coefficient for pancreatic segmentation was 0.746 ± 0.099, which is lower than previously reported values. This may be attributable, in part, to imaging artifacts such as streak artifacts caused by arm positioning. This limitation may have affected the accuracy of the estimated pancreatic volumes; however, the overall patterns were consistent with those reported in previous studies. Finally, although clinically significant abnormalities were excluded, lesions not considered clinically significant, such as simple cysts, were not controlled for and may have influenced the estimated organ volumes.

In conclusion, longitudinal low-dose CT-based volumetric analysis characterized nonlinear age-related trajectories in abdominal organ volumes across multiple organs. Liver, pancreas, and kidney volumes showed a mild increase or plateau at approximately 40–60 years of age, followed by a gradual decline, whereas splenic volume showed a progressive decrease. Analysis of the first derivatives further demonstrated age-dependent transitions in volume change rates, with differences in timing and magnitude between sexes. These findings provide a comprehensive characterization of age-related organ volume changes and may serve as a reference for their interpretation in clinical and research settings.

## Data Availability

The original clinical data cannot be publicly shared because of ethical and privacy restrictions.

## Acknowledgements

The authors would like to thank Dr. Naoto Hayashi for manual annotation of abdominal organs used for segmentation performance evaluation. The Department of Computational Diagnostic Radiology and Preventive Medicine, The University of Tokyo Hospital, is sponsored by HIMEDIC Inc. and Siemens Healthcare K.K.

## Compliance with Ethical Standards

### Guarantor

The scientific guarantor of this publication is Takeharu Yoshikawa.

### Conflict of interest

The authors of this manuscript declare no relationships with any companies, whose products or services may be related to the subject matter of the article.

### Statistics and Biometry

One of the authors has significant statistical expertise.

### Informed consent

Written informed consent for use of imaging data in research was obtained at the time of the whole-body medical screening program.

### Ethical approval

Institutional Review Board approval was obtained.

### Study subjects or cohorts overlap

This study used data from a previously described cohort [1]. Although this cohort has been used in multiple prior studies, different inclusion criteria and a distinct research objective were applied in the present analysis.

### Methodology

- retrospective
- observational
- performed at one institution

## Notes

### Competing Interest Statement

The authors have declared no competing interest.

### Funding Statement

This study did not receive any funding.

### Author Declarations

Research Ethics Committee of the Faculty of Medicine of the University of Tokyo gave ethical approval for this work.

