## Supplemental material for "Age-related nonlinear trajectories of abdominal organ volumes on CT: a longitudinal study"

### Supplementary Material

#### S1. Segmentation performance evaluation

For the evaluation of segmentation performance, subjects who underwent their first screening between February 10, 2015, and March 31, 2015, were identified (n = 75). One subject with kidney disease was excluded, leaving 74 eligible subjects. From these, 40 subjects were randomly selected to construct the evaluation dataset. Each target organ was manually delineated by a board-certified radiologist (N.H., with 35 years of experience in CT interpretation) using voxel-wise annotation. The annotations were performed using the web-based image database system CIRCUS DB [20] and used as reference standards for performance evaluation.

The segmentation performance of TotalSegmentator was evaluated by comparing the automated segmentation results with the reference standards. For each organ, volumetric measurements obtained from the automated method and reference standard were compared. The error ratio (%) was calculated as the absolute difference between the automated and manual volumes divided by the manual volume. Segmentation accuracy was further assessed using the Dice similarity coefficient (Dice), which measures the spatial overlap between the automated segmentation and the reference standard. The results are summarized in Table S1.

**Table S1.** Segmentation performance of TotalSegmentator for each organ.

| Organ | Volume (auto) [ml] | Volume (manual) [ml] | Error ratio [%] | Dice |
|---|---|---|---|---|
| Liver | 1366.9 ± 292.7 | 1328.8 ± 295.1 | 3.6 ± 1.7 | 0.957 ± 0.006 |
| Spleen | 125.2 ± 49.6 | 120.1 ± 49.8 | 6.7 ± 5.7 | 0.917 ± 0.027 |
| Pancreas | 71.5 ± 22.2 | 70.2 ± 20.8 | 19.9 ± 15.6 | 0.746 ± 0.099 |
| Right kidney | 145.7 ± 37.3 | 140.9 ± 37.0 | 6.9 ± 4.5 | 0.917 ± 0.033 |
| Left kidney | 156.0 ± 43.3 | 153.2 ± 41.1 | 5.3 ± 4.9 | 0.919 ± 0.024 |

Data are presented as mean ± standard deviation.

#### S2. Outlier removal

Outlier removal was performed based on the distribution of organ volumes derived from automated segmentation using a robust statistical framework adapted from a previous study [11]. In this study, only the distribution-based filtering step was applied. For each organ, volumetric measurements derived from automated segmentation were first log-transformed using log1p to stabilize variance. Robust summary statistics, including the median and median absolute deviation (MAD), were then calculated. Robust z-scores were computed as the deviation from the median scaled by the MAD, and values with an absolute robust z-score greater than 4 were identified as outliers and excluded. This procedure was applied consistently across all organs.

#### S3. Analysis of the baseline age–time model

The baseline age–time model described in the previous study [11] was adapted to the present dataset. Terms related to scanner manufacturer and contrast agent use were omitted, as all examinations were performed using a single scanner model and without contrast enhancement. In addition, fractional polynomial (FP) powers for baseline age were re-estimated using the current dataset, and the selected FP terms for each organ are summarized in Table S2. Only the mean structure ($\mu$) was modeled using FPs, consistent with their use in the longitudinal model in the previous study. This approach ensures that the model structure is consistent with the previous study [11] while allowing appropriate adaptation to the characteristics of the present cohort. FP powers were selected from the standard set {−2, −1, −0.5, 0, 0.5, 1, 2, 3} by optimizing the Bayesian information criterion (BIC), consistent with the approach used in the previous study [11].

**Table S2.** Selected fractional polynomial (FP) for each organ.

| Organ | Selected FP |
|---|---|
| Liver | $\beta_1 \cdot {age_b}^{-2} + \beta_2 \cdot {age_b}^{-2}\log(age_b)$ |
| Spleen | $\beta_1 \cdot {age_b}^{2} + \beta_2 \cdot {age_b}^{3}$ |
| Pancreas | $\beta_1 \cdot {age_b}^{-2} + \beta_2 \cdot {age_b}^{0.5}$ |
| Right kidney | $\beta_1 \cdot {age_b}^{3} + \beta_2 \cdot {age_b}^{3}\log(age_b)$ |
| Left kidney | $\beta_1 \cdot {age_b}^{3} + \beta_2 \cdot {age_b}^{3}\log(age_b)$ |

Fig. S1 shows age-related changes in organ volume based on the baseline age–time model across all examined organs. In both sexes, liver, pancreas, and kidney volumes showed a mild increase or plateau in midlife, followed by a gradual decline with increasing age, whereas splenic volume showed a progressive decrease across the age range. Overall, similar patterns were observed between males and females, with consistently larger organ volumes in males.

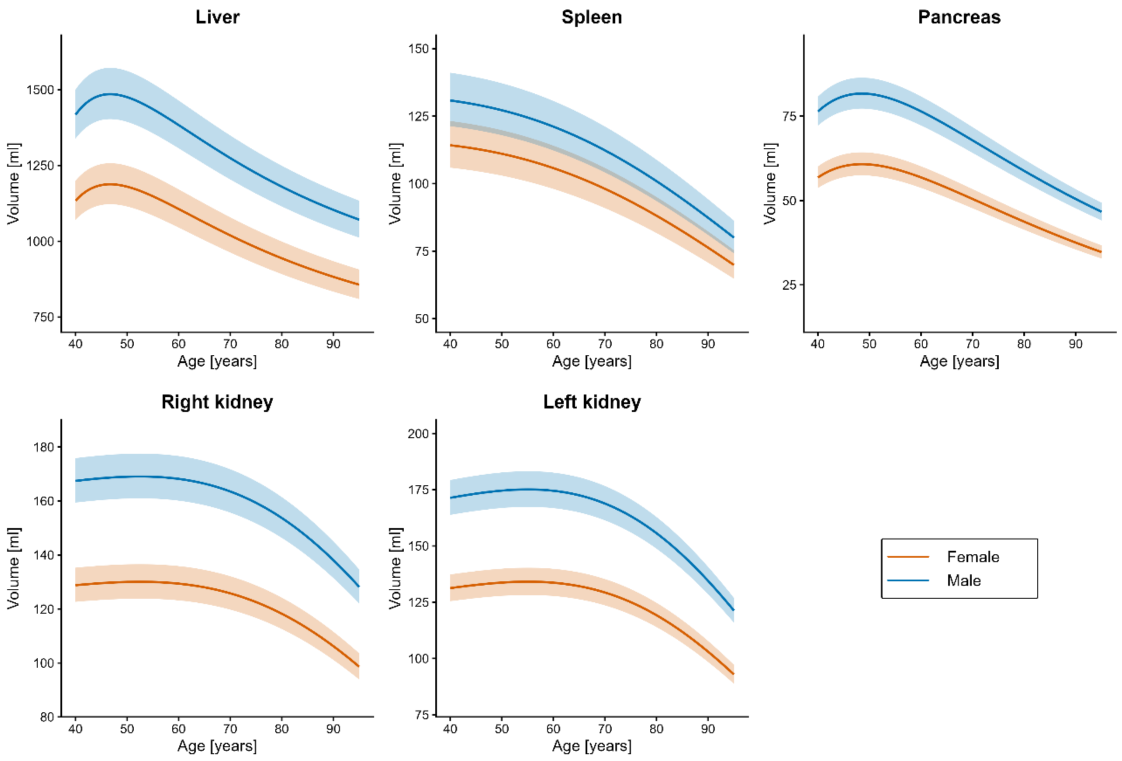


**Fig. S1.** Visualization of generalized additive mixed model fits for organ volumes based on the baseline age–time model. Curves show estimated organ volumes as a function of baseline age for female participants (red) and male participants (blue). Shaded ribbons indicate model-based 25th–75th percentile bands.

#### S4. Distribution of examination intervals

The distribution of intervals between consecutive CT examinations is shown in Fig. S2. The median interval was 11 months (interquartile range [IQR], 11–12 months). Most intervals were observed around 11–12 months, with a small proportion of longer intervals.

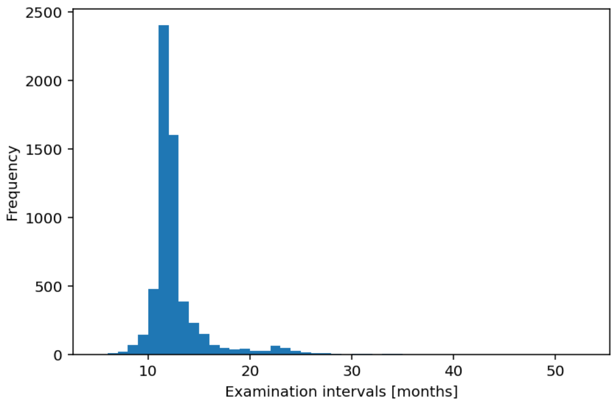


**Fig. S2.** Distribution of intervals between consecutive CT examinations.
